# Genetic exploration of the relationship between liability to psychiatric disorders and acne vulgaris

**DOI:** 10.1101/2025.06.09.25329292

**Authors:** Brittany L. Mitchell, Michelle K. Lupton, Miguel E. Rentería, Michael A. Simpson, William R. Reay

## Abstract

**Background:** Observational epidemiology suggests a link between the dermatological disorder acne vulgaris and several psychiatric disorders. However, the biological mechanisms that underlie the relationship between acne and mental health are poorly characterised.

**Methods:** Here, we employed a genetic approach using large-scale genome-wide association studies of acne and ten psychiatric disorders to both estimate causal effects and uncover potential shared genetic risk factors.

**Results:** While multiple psychiatric disorders displayed evidence of small-to-moderate genetic correlations with acne, only genetic liability to schizophrenia was associated with a putative causal effect on the risk of acne. We then investigated shared genetic risk architecture between schizophrenia and acne using a Bayesian approach. This revealed a subset of genetic loci associated with both acne and schizophrenia via shared or different causal variants, implicating biological processes including glutamatergic signalling. Finally, we found that genetic risk for schizophrenia was also associated with increased acne severity.

**Conclusions:** In summary, we revealed genetic support for a biological relationship between acne and schizophrenia that may at least partially drive the elevated rates of acne amongst people living with schizophrenia.

## INTRODUCTION

Acne vulgaris, the most common skin disorder, is characterized by inflammation of the pilosebaceous unit, resulting in lesions such as comedones, papules, pustules, nodules, and cysts. Lesions typically appear on the face, neck, chest, and back, commonly beginning during puberty. Acne affects over 85% of adolescents, with approximately 10% experiencing moderate to severe cases, and around 8% developing severe forms that often persist into adulthood (1,2).

Beyond physical impacts, acne significantly affects mental health. Severe acne is associated with heightened stress, social withdrawal (3–5), depression, anxiety, poor self-image, and suicidal tendencies. Individuals with severe acne are up to three times more likely to experience suicidal ideation and other psychiatric disorders than those with little or no acne (4). Acne accounts for an estimated 5 million Disability-Adjusted Life Years (DALYs) globally, highlighting its substantial psychological and societal burden (6).

The relationship between acne and mental health disorders may be bidirectional. Acne’s visible and chronic nature contributes to psychological distress, whereas stress itself can exacerbate acne by increasing androgen and cortisol levels, promoting inflammation and sebum production (7,8). Emerging evidence also suggests deeper biological connections, involving neurotransmitter systems such as serotonin and dopamine (9,10), systemic inflammation pathways (11), and retinoic acid signalling, which is a critical regulator in immune function, skin health, and neurodevelopment (12,13).

Despite these biological links, the causal relationship between acne and mental health disorders remains unclear. Most existing studies are observational, making it challenging to determine if acne contributes to mental health symptoms, if underlying biological factors predispose individuals to both conditions, or if a combination occurs. Both acne and psychiatric conditions are significantly heritable (2,14). Recently, large genome-wide association studies (GWAS) revealed genetic correlations between acne and psychiatric disorders, including depression, bipolar disorder and schizophrenia (15). However, the causal nature of these associations and specific biological pathways involved remain uncertain.

Here, we leverage recent GWAS data on acne and ten psychiatric disorders to estimate genetic correlations, assess causality, and identify shared genetic architecture. By biologically annotating these links, we aim to uncover mechanisms underlying acne and psychiatric conditions, with potential implications for targeted therapies.

## MATERIALS AND METHODS

### Genome-wide association study data

GWAS summary data are listed in Table 1, with full details available in the original publications referenced. Psychiatric phenotypes included attention-deficit/hyperactivity disorder (ADHD), anorexia nervosa (AN), autism spectrum disorder (ASD), bipolar disorder (BIP), generalised anxiety disorder (GAD), major depressive disorder (MDD), obsessive compulsive disorder (OCD), post-traumatic stress disorder (PTSD), schizophrenia (SZ), and Tourette’s syndrome (TS) (16–25).

**Table 1.** Primary genome-wide association study datasets used in this study.

| Phenotype | N<sub>Cases</sub><sup>1</sup> | N<sub>Controls</sub> | Reference |
| --- | --- | --- | --- |
| Attention-deficit hyperactivity disorder (ADHD) | 38691 | 186843 | (16) |
| Anorexia nervosa (AN) | 16992 | 55525 | (17) |
| Autism spectrum disorder (ASD) | 18381 | 27969 | (18) |
| Bipolar disorder (BIP) | 41917 | 371549 | (19) |
| Generalised anxiety disorder (GAD) | 31977 | 82114 | (24) |
| Major depressive disorder (MDD)<sup>2</sup> | 294322 | 741438 | (25) |
| Obsessive-compulsive disorder (OCD) | 2688 | 7037 | (23) |
| Post-traumatic stress disorder (PTSD) | 137136 | 1085746 | (20) |
| Schizophrenia (SZ) | 53386 | 77258 | (21) |
| Tourette's syndrome (TS) | 4819 | 9488 | (22) |
| Acne vulgaris (acne) | 20165 | 595231 | (15) |

<sup>1</sup>Sample sizes are the maximum for a GWAS and may vary across variants
<sup>2</sup>A subset of the primary depression GWAS meta-analysis of samples of European ancestry, excluding 23andMe (cases and controls)

We further examined the larger trans-ancestral GWAS of SZ from the same publication as a sensitivity analysis, given that SZ was the trait that displayed the most pervasive relationship with acne in the subsequent components of the study (74,776 cases, 101,023 controls, ∼26% non-European) (21).

### Genetic correlation and latent causal variable analyses

Genetic correlation (*r*_g_) between acne and each psychiatric disorder was first estimated using linkage disequilibrium (LD) score regression (LDSR) (26,27) (Supplementary Methods). We also compared correlation estimates for traits that surpassed Bonferroni multiple-testing correction to an alternate genetic correlation estimator, the High-Definition Likelihood (HDL) method (v1.4.0) using a UK Biobank LD reference panel (https://github.com/zhenin/HDL) (28). For genetically correlated trait pairings, we then constructed a latent causal variable (LCV) model (29), as described elsewhere (30). Briefly, this model is used to estimate partial genetic causality and leverages the effect of the panel of ‘munged’ HapMap3 variants genome-wide on either trait to evaluate evidence whether the effect of one trait on the second is larger than in the reverse direction (Supplementary Methods).

### Mendelian randomisation

Mendelian randomisation (MR) approaches were then implemented to further explore putative causal relationships between the genetic liability of acne and each of the ten psychiatric disorders (Supplementary Methods) (31,32). We note that given both our exposure and outcome traits are binary this does present some methodological challenges in terms of the effect size estimate (33). As a result, we focus our interpretation on the directionality and strength of evidence to reject the null hypothesis in these models rather than the magnitude of any estimates.

The primary MR model we used in this study was “Causal Analysis Using Summary Effect Estimates” (CAUSE) as implemented using the CAUSE R package v1.2.0, which is also able to account for sample overlap (34). This is a more polygenic approach than most traditional MR implementations as it seeks to compare the fit of a model that assumes that the observed association between the exposure and outcome arises from horizontal pleiotropy versus a model that allows both horizontal pleiotropy and a causal effect. Model comparison was performed using the expected log pointwise posterior density (ELPD) to test whether the ‘causal’ inclusive model fits better than the ‘sharing’ only model. In line with the original CAUSE publication, ∼ 1 million random variants from the GWAS were used to estimate nuisance parameters in the model, followed by specifically using a set of clumped variants that are at least nominally associated with the exposure to perform model comparison (*P*□<□0.001, *r*^2^□<□0.01, 1000 genomes phase 3 European reference panel). Another key aspect of the CAUSE framework is that it assumes a proportion of variants (denoted *q*) exhibit what is termed ‘correlated pleiotropy’ whereby variants effects on the exposure and outcome are correlated through a shared heritable factor (e.g., a shared pathway), whilst the remaining variants are correlated arising from different factors (uncorrelated pleiotropy). We describe the modelling considerations regarding the beta prior on *q* in the Supplementary Methods.

We also conducted sensitivity analyses using several more ‘traditional’ MR methods which use a subset of strongly exposure-associated genetic variants as instrumental variables (IVs) using the TwoSampleMR R package (Supplementary Methods) (32,35). Given that the variants associated with the exposure that are used in MR are heterogenous, we hypothesised that particular subsets of genetic instruments may have more consistent effects - potentially through shared biological mechanisms. We explored this for the bidirectional Acne/SZ models by leveraging the method MR-Clust (v0.1.0) (36). Briefly, MR-Clust is a mixture model framework that aims to identify latent clusters of genetic instruments with similar exposure-outcome effects (Supplementary Methods).

### Pairwise analysis of genome-wide association study data

We used the Bayesian approach pairwise GWAS (PW-GWAS, https://github.com/joepickrell/gwas-pw) as our primary method to identify signals shared between schizophrenia and acne that may be driven by the same underlying causal variant, as described previously (37,38). This method jointly models the association of two phenotypes using 1,703 independent genomic segments (LD blocks) (39). For each LD block, and its bivariate effect size distribution, the prior probability and posterior probability (PPA) is estimated under four competing models, with models 1 and 2 referring to associations only seen for one of the two traits, respectively, whilst model 3 is a region that contains a shared causal variant that influences both traits, our primary model of interest. The fourth model relates to regions associated with both traits but influenced by independent causal variants. We used a PPA for model 3 threshold of 80% to define regions (LD blocks) with a shared causal variant, and a threshold of 50% as suggestive, although we acknowledge that these values are somewhat arbitrary. We visualised these signals using an ideogram generated by http://visualization.ritchielab.org/phenograms/plot. We also investigated variants that may underlie these associations and compared them to another Bayesian colocalisation approach (coloc) (40), as outlined in the supplementary methods. For putative shared risk genes, a high-confidence protein-protein interaction network was constructed for that gene using STRING version 11, in line with previous work (41), and overrepresentation in biological pathways tested using g:Profiler (42).

### The association of high confidence schizophrenia risk genes with acne

We also used a more granular approach to detect shared risk signals between SZ and acne. We extracted the association from the acne GWAS for 12 high-confidence SZ risk genes that were supported by at least two lines of evidence from the following in the latest GWAS publication from the Psychiatric Genomics Consortium: i) finemapping, ii) summary-based Mendelian randomisation expression prioritisation, and iii) evidence for rare variant association with schizophrenia or a neurodevelopmental disorder - as outlined elsewhere (21). Shared SZ-acne association signals were visualised using the R packages topr v.2.0.1 and locuscomparer v1.0.0 (43,44).

### Modelling the association between schizophrenia genetic risk and acne severity

To investigate the potential relationship between schizophrenia (SZ) genetic risk and acne severity, we first created polygenic scores (PRS) for SZ using SBayesR. SBayesR was applied to summary-level data from a large GWAS meta-analysis of schizophrenia, resulting in PRS that capture genetic liability across the entire genome. Next, we tested the association of these SZ PRS with acne severity across different groups in an independent sample, the Prospective Imaging Study of Ageing (PISA) sample (45), as we have outlined previously (15). Acne severity was categorized into groups based on self-reported assessment on a four-point scale that ranged from none, mild, moderate, to severe acne (N=1,894 unrelated individuals). Pairwise comparisons between acne severity categories and the reference group (“None”) were assessed using a linear model in R, with the acne severity grouping entered as a factor and “None” set as the reference level.

## RESULTS

### Positive genetic correlations observed between acne vulgaris and psychiatric disorders

We replicated the observation of non-zero acne-psychiatric genetic correlations from our previous work (15), with larger psychiatric GWAS tested herein (Table 2). Significant positive genetic correlations after Bonferroni correction (*P* < 5×10^−3^, ten traits tested) were estimated with SZ, OCD, BIP, and MDD. A nominally significant (*P* < 0.05) positive correlation was also found with PTSD, whilst the estimate with GAD was a non-significant trend (*P* = 0.052). The most statistically significant estimate of genetic correlation with acne was found with SZ (*r*_g_ = 0.14, *SE* = 0.028, *P* = 4.2 × 10^−7^). We then tested the consistency of these findings using an alternate r_g_ estimator (HDL method) and a modified weighting scheme for LDSR in the LCV R package, yielding similar results (Supplementary Table 1). Taken together, this further supports the notion of shared genetic liability between acne and psychiatric disorders, albeit with small to moderate effect sizes.

**Table 2.**
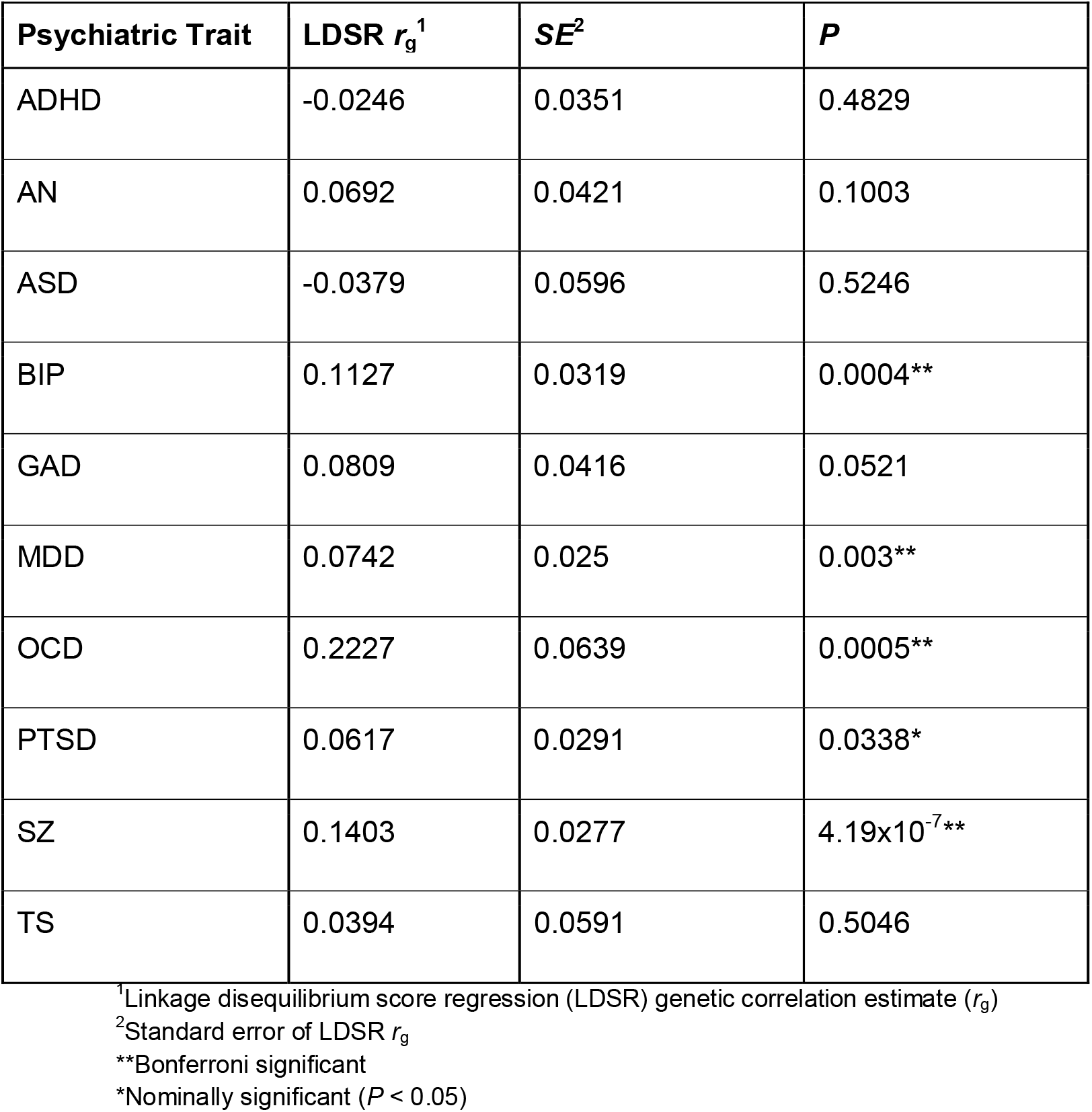
Estimated genetic correlations between psychiatric disorders and acne vulgaris (LDSR)

### Evidence for an effect of genetic liability to schizophrenia on increased risk of acne vulgaris

We used latent causal variable (LCV) models to estimate the potential direction of causality between acne liability and psychiatric disorders for genetically correlated pairs. There was no strong evidence of partial genetic causality (non-zero |GCP| > 0.6) found between any acne-psychiatric trait pairs (Supplementary Table 2). These LCV modelling results would suggest that the observed genetic correlations reflect horizontal pleiotropy or other mechanisms rather than a direct relationship between liability for acne and psychiatric disorders. However, the LCV approach only models a shared latent factor and how genetic risk for either trait loads onto that factor. Therefore, we attempted to distinguish between correlated pleiotropy and causal effects between acne and psychiatric genetic liability using CAUSE (Figure 1A, Supplementary Tables 3,4). We found that modelling a causal effect of genetic liability to SZ on acne risk fitted better than a model of a null relationship (ΔELPD_NullvsCausal_ = -10.41, *SE* = 4.34, *P* = 0.017) or that of a ‘sharing’ (correlated pleiotropy) only relationship (ΔELPDS_haringvsCausal_ = -3.85, *SE* = 1.64, *P* = 0.019). The posterior median estimate of the causal term was positive, indicating that increased genetic liability to schizophrenia was associated with increased odds of acne. This was broadly consistent using different priors, although the sharing-only and causal-inclusive models were not significantly distinguishable using the more conservative prior *q ∼ Beta*(1,2) (Figure 1A, Supplementary Table 5). There was some directionally consistent evidence also seen in the other direction (acne liability → SZ), although the differences between the sharing and causal model were not statistically significant. Very similar results were obtained using the trans-ancestral SZ GWAS (Supplementary Table 5). Taken together, these data suggest that increasing genetic liability to schizophrenia may, in turn, increase liability to acne vulgaris, although we highlight that the precision of the ΔELPD is relatively low, as reflected by somewhat nominal statistical significance for model comparison.

**Figure 1.**
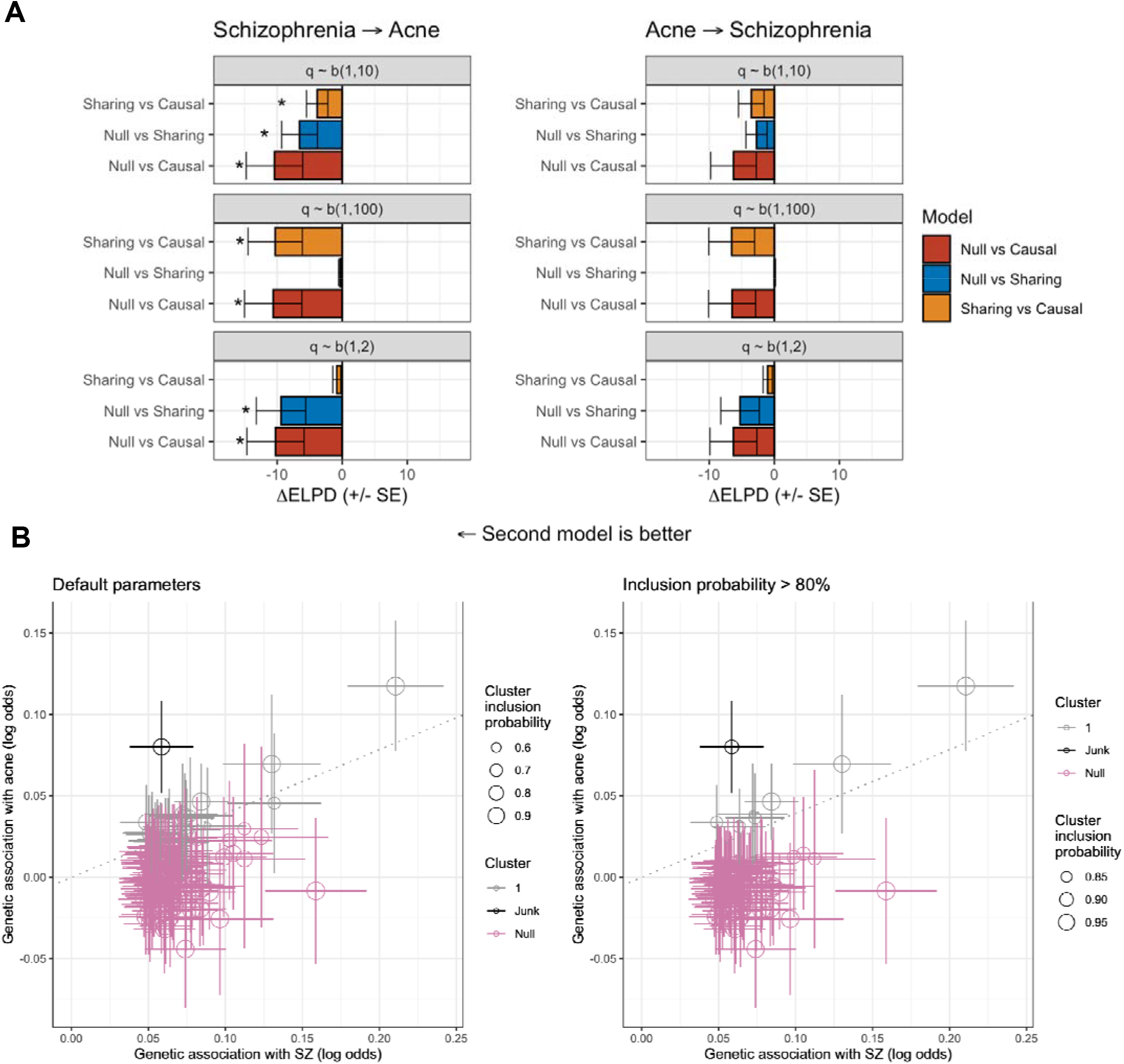
Modelling the relationship between genetic liability to schizophrenia and acne vulgaris. (**A**) The CAUSE modelling framework was utilised to estimate the effect of genetic liability to schizophrenia on acne vulgaris (left), and conversely, genetic liability to acne vulgaris on schizophrenia (right). In both instances, the change in expected logwise point density (.t1ELPD) is plotted, with error bars denoting the standard error, between the two models in three configurations: null versus sharing, null versus causal, and sharing versus causal. Three different beta priors were implemented: q ∼ Beta(1,10), q ∼ Beta(1,2), and q ∼ Beta(1,100). An asterisk denotes a significant difference in model fit (P < 0.05). (**B**) The results of the MR-clust approach are visualised which investigated the association of individual schizophrenia IVs with acne. This is a mixture model framework that seeks to identify clusters of IVs with similar causal estimates. The size of the point denotes the cluster inclusion probability, relating to the conditional probability of cluster membership. The null cluster, coloured pink, relates to IVs with null effect, whilst the black “junk cluster” includes variants that were not parsimoniously assigned to any cluster. The cluster of interest is denoted in grey, with a trend line indicative of the mean cluster effect. The leftmost panel utilised default parameters for cluster assignment, whilst the right panel only retained variants with an inclusion probability > 80%.

In line with our CAUSE results, conventional IV-driven bidirectional Mendelian randomization (MR) found strongest evidence for a causal relationship between SZ liability to acne (β = 0.093, *P* = 0.0002), with no significant evidence for a causal relationship in the other direction (β = 0.067, *P* = 0.058) (Supplementary Table 6). Sensitivity analyses using the weighted median, weighted mode, and Egger regression methods were generally consistent with the IVW results. No strong evidence of a causal effect was observed from acne to other psychiatric conditions, with only a nominally significant relationship between acne and autism (*β* = 0.055, *P* = 0.034) in the IVW analysis, but none of the other MR methods. In contrast to the CAUSE modelling, we also observed a significant estimate for a causal relationship of genetic liability to bipolar disorder on the risk of acne (*β* = 0.122, *P* = 0.0019). However, given the lack of consistent evidence across the multiple MR methods and wide confidence intervals, these findings should be interpreted with caution.

Finally, we investigated how SZ-associated genetic signals used as IVs for MR modelling may cluster together in terms of their association with acne liability (Figure 1B). We found that there was a subset of SZ IVs that clustered together and had a positive cluster mean, that is, increased genetic liability to SZ associated with increased liability to acne, whilst most IVs were in the ‘null’ cluster. These results were generally consistent using a posterior inclusion probability > 80% for cluster assignment (Figure 1B) and the trans-ancestral SZ GWAS (Supplementary Figure 1). We explored the potential biological significance of the variants that comprise cluster 1 by assigning each variant to a gene based on physical position and exploring the biological overrepresentation of genes. This revealed a significant (FDR < 0.05, > 2 intersecting genes) enrichment of cluster 1 signals amongst the ‘*synapse*’ and ‘*glutamatergic synapse*’ gene-ontology cellular component sets.

### Exploring the genetic overlap between schizophrenia and acne vulgaris

Using PW-GWAS, we found 8 independent LD blocks with evidence for a shared variant signal that influences both SZ and acne liability using a strict posterior probability of association (*PPA*) threshold of 80%, with an additional 13 such LD blocks detected using *PPA* ≥ 50% (Figure 2, Supplementary Table 7). The majority of the LD blocks implicated with a strong shared variant signal were in or proximity to the extended MHC region (5 of 8), which is unsurprising given the strong association of that locus with both SZ and acne. We also found LD blocks more aligned with the model of an association with both traits driven by different causal variants (model 4), with 13 LD blocks exhibiting strong support for this model (*PPA* ≥ 80%). Outside the MHC, the 90% credible sets from the third PW-GWAS model (shared causal variant) identified three LD blocks each with a single variant having *PPA* > 30% (Figure 3B-C). Mapping these variants to their closest gene based on physical proximity prioritised three genes that may represent shared signals between SZ and acne - *DLG1, PCNXL3*, and *MTIF3*. We also identified two additional potential shared genes by selecting credible sets without a single high-PPA variant, but where all variants mapped to the same gene, revealing two further potential shared protein-coding genes - *SCAPER* and *ZSWIM6*. A high-confidence protein-protein interaction network for *DLG1* was strongly enriched for several pathways, including those related to glutamatergic neurotransmission (Supplementary Table 8), which is logical given the role *DLG1* is purported to play in synaptic scaffolding (46). Interestingly, this signal mapped to *DLG1*, which we previously identified as genome-wide significant in the acne GWAS, has a discordant direction of effect between schizophrenia and acne. This highlights that, whilst overall the genetic relationship between acne and schizophrenia is that of a positive correlation, individual signals can diverge from this, like *DLG1*. Finally, we compared the PW-GWAS results to those of another Bayesian method, ‘coloc’ (Supplementary Figure 2). Overall, the most confident regions for the third PW-GWAS model tended to also support the analogous coloc prior hypothesis of a shared causal variant, although there was more discordance between lower PPA regions, where some more strongly supported the coloc hypothesis of distinct causal variants.

**Figure 2.**
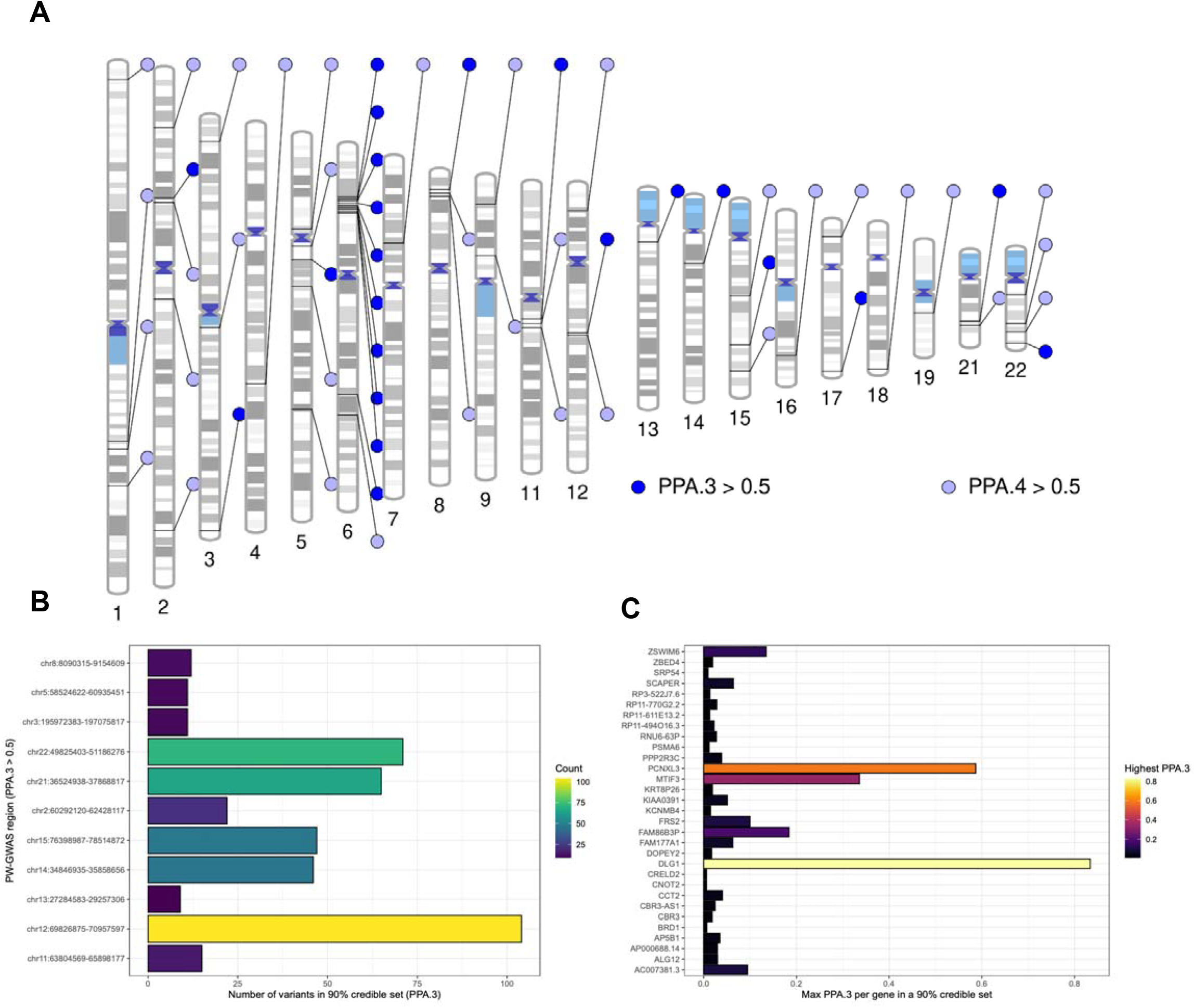
The shared genetic architecture of schizophrenia and acne. (**A**) Ideogram of LD blocks which are estimated to have some shared genetic relationship between schizophrenia and acne using a threshold of a posterior probability (PPA) > 50% for either model 3 (PPA.3 > 0.5), which assumes a single shared causal variant, or model 4 (PPA.4 > 0.5), which assumes a different causal variant for each phenotype in the LD block. (**B**) 90% credible set of variants in LD blocks with PPA.3 > 0.5 given the variant level PPA that they are the shared causal variant, with the number of variants in each credible set plotted. (**C**) Genes that may be impacted by these credible set variants (based on the closest gene), with the highest variant level PPA.3 per gene plotted.

**Figure 3.**
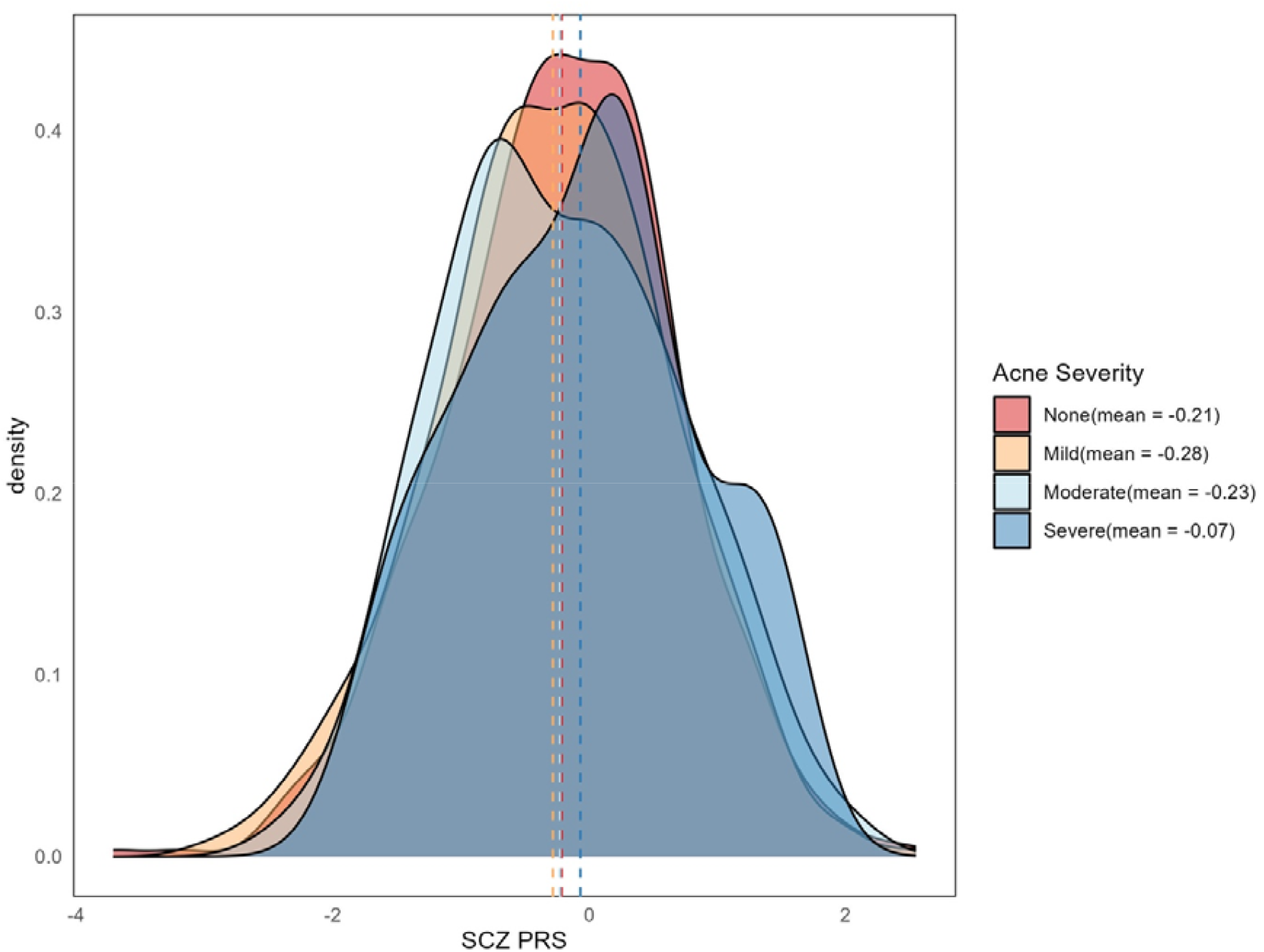
Distribution of schizophrenia polygenic risk scores (SCZ PRS) by acne severity in the PISA sample (N=1,894). Kernel density plots depict the distribution of SCZ PRS across four acne severity categories: None, Mild, Moderate, and Severe. Vertical dashed lines indicate the mean PRS for each group, with corresponding values provided in the legend.

In addition, to the region-based approach (LD blocks) described above, we also screened eight genes associated with SZ that represented the most confidently prioritised genes from the latest SZ GWAS for their association with acne (*BCL11B, CACNA1C, GRIN2A, KANSL1 RERE, SLC39A8, SP4, CUL9, FURIN, LINC00320, SNAP91, ZNF823*). There was a signal for acne mapped based on physical location within the *RERE* gene, an important regulator of retinoic acid signalling, that approached genome-wide significance (Supplementary Figure 3). However, visual inspection of the *RERE* signal for SZ and acne suggested that a distinct causal variant may underlie this (line with the PW-GWAS results for that LD block), necessitating additional exploration of this region.

### Schizophrenia genetic risk is associated with acne severity

We examined whether SZ PRS differed according to acne severity in the PISA sample. As shown in Figure 3, the distribution of SZ PRS varied across acne severity categories. Individuals reporting severe acne had higher average SZ PRS (mean = -0.07) compared to those reporting no acne (mean = -0.21), with a progressive shift in distribution evident from mild to severe cases. This shift was nominally statistically significant (*P* = 0.042). While the overall effect sizes were modest, this pattern suggests a potential association between greater schizophrenia genetic liability and increased acne severity.

## DISCUSSION

There is increasing recognition of shared biological mechanisms between acne and mental health disorders. Studies, including genome-wide association studies (GWAS), have begun to uncover genetic overlap, suggesting that common pathways, such as inflammatory processes or hormonal imbalances, may contribute to both conditions (15). By investigating the connection between mental health traits and acne, we aim to better understand the underlying biological aetiology, which could ultimately inform the development of more effective and targeted treatments. Improved therapeutic approaches that address both the psychological and dermatological aspects of acne could enhance overall patient outcomes and quality of life, reducing the burden of both the skin condition and its associated mental health challenges.

In this study, we built on previous evidence to reveal loci that are putative shared genetic risk factors between acne and schizophrenia, as well as some evidence to suggest that increased genetic liability to schizophrenia may directly be associated with a small-effect size increase in the risk of acne. These data broadly support existing observational epidemiological evidence studying the relationship between the two disorders. For instance, retrospective analysis of a decade of data from the United States National Inpatient Survey, which captures a representative subset of hospitalisations, revealed individuals living with acne were more likely to be diagnosed and hospitalised with schizophrenia along with several other psychiatric diagnoses, such as depression, anxiety, and substance use disorders (47). Moreover, there is emerging evidence for a role of dopaminergic signalling in the skin (48,49), raising the possibility that dopaminergic dysfunction attributed to schizophrenia could have dermatological consequences. Dissection of plausible shared risk genes from the PW-GWAS modelling revealed specific candidates that could be subjected to further study - including *DLG1, PCNXL3*, and *MTIF3*. Overrepresentation analysis of the cluster of the closest genes to schizophrenia risk loci, more specifically related to acne liability (MR-Clust analyses), also broadly supported a role for glutamatergic signalling in the relationship between the two disorders. However, we acknowledge the limitation that these genes were inferred using the metric of the genes closest based on physical genomic position, which may not always be the underlying causal gene. Whilst the significance of glutamatergic signalling with respect to schizophrenia has been well established, the role of glutamate in the skin and periphery may also be directly relevant to acne. Recent rodent data suggest that cutaneous sensory neurons may contribute to skin homeostasis via glutamate release that, in turn, downregulates inflammatory mast cell activation (50). The influence of specific schizophrenia risk variants linked to glutamate and glutamatergic neurotransmission could be further studied in the context of peripheral cutaneous neuronal activity, as well as inflammatory mediators linked to acne pathogenesis. Finally, we highlight that the region spanning the strongly supported common-variant schizophrenia risk gene *RERE* was also associated with acne, albeit likely via a different causal variant. This finding is potentially notable in the context of the relationship between *RERE* and the regulation of retinoic acid (51), with retinoic acid isoforms and related synthetic derivatives commonly indicated as topical acne treatments. There has been significant controversy over the link between retinoids indicated for acne, like isotretinoin (13-*cis* retinoic acid) and psychiatric outcomes, with some evidence to suggest that topical retinoids are associated with increased risk of affective disorders and suicidality (52). However, more recent data have disputed this and found that isotretinoin may exert protective effects on mental health (53). Shared genetic risk loci between schizophrenia and acne broadly supports existing clinical trial and genetic data, which implicates altered retinoic acid signalling in its pathogenesis (12), although additional work is needed to further understand this. In general, genetics may be a useful tool for studying the impact of retinoic acid signalling on the brain and psychiatric outcomes, given that causal inference to resolve the relationship between topical retinoids (e.g., isotretinoin) and mental health has not previously been feasible.

Our analysis revealed no direct causal effect of the genetic liability of acne on depression, nor *vice versa*. While we identified some evidence of shared genetic architecture between the two conditions, indicating potential underlying biological connections, no evidence for a direct causal relationship emerged. These null results suggest that while acne and depression may share common genetic influences, such as inflammatory processes, hormonal regulation, and immune system dysfunction, acne and depression may not directly drive one another. Rather, they may interact in more complex, indirect ways that are yet to be fully understood. The shared pathways we observed could also involve mechanisms, including altered skin barrier function or stress-related pathways that affect both mental and dermatological health (54). Further research is needed to further explore these shared pathways, using more refined methodologies, larger sample sizes, and longitudinal data, to better understand the intricate interactions between these traits.

Finally, it is important to acknowledge that while our genetic study design has methodological advantages, it also has some key limitations that must be considered when interpreting these results. Firstly, GWAS of common variants capture only part of the genetic architecture, future work should consider pleiotropy between acne and mental health from rare and structural variants. Secondly, genetic correlations may reflect shared biology or arise from an unmeasured trait influencing both conditions (27). Our identification of variants contributing to both acne and schizophrenia suggests some shared biological basis, but it does not rule out the role of other intermediate or confounding phenotypes. Thirdly, there are also several important limitations to the use of genetic variants for causal inference, particularly for binary traits, as outlined extensively elsewhere (30–33). As a result, we caution that our data, which reports a direct effect of liability to schizophrenia, indexed by genetics, on acne, requires further replication and validation using other epidemiological methods. Finally, it also remains unclear how specific our findings are to acne, versus more generalised shared genetic risk architecture that may exist between psychiatric phenotypes and other related dermatological conditions. Our findings are also limited to European ancestry populations, which in turn also decreases the generalisability of the data. In future, we believe that building on our findings may reveal important insights into both acne and mental health, such as the impact that potential shared aetiological influences (e.g., neurotransmission) may exert on the skin and central nervous system, respectively.

## Data Availability

All data utilised in this study is publicly available

https://pgc.unc.edu/

https://www.ebi.ac.uk/gwas/studies/GCST90092000

https://ipsych.dk/en/research/downloads

## ACKNOWLEDGEMENTS

W.R.R and BLM were supported by an Australian National Health and Medical Research Council (NHMRC) EL1 Investigator Grant fellowship (2025671 and 2017176, respectively). This research was conducted with the support of the Australian Skin and Skin Cancer Research Centre through the ASSC Early Career Researcher Grant. M.E.R. thanks the support from the Rebecca L. Cooper Medical Research Foundation through an Al & Val Rosenstrauss fellowship (F20231230). The Prospective Imaging Study of Ageing (PISA) was funded by an NHMRC Dementia Research Team Grant [APP1095227]. MAS is supported by the Medical Research Foundation (MRF-RGL-ASD-23-108) and the Leo Foundation (LF-OC-22-001033)

## DISCLOSURES

The authors declare no competing financial interests or conflicts of interest.

## Notes

### Competing Interest Statement

The authors have declared no competing interest.

### Author Declarations

Ethics committee of the University of Tasmania waived ethical approval for this work

