## Supplementary Text and Figures for "Genetic exploration of the relationship between liability to psychiatric disorders and acne vulgaris"

**Supplementary Methods and Figures**

### SUPPLEMENTARY METHODS

#### Genetic correlation

LDSR identifies trait pairs for which genetic effects exhibit a non-zero correlation genome-wide, with variants that exhibit more LD with neighbouring loci (higher LD scores) upweighted as these variants are posited to represent true causal signals. The primary LDSR analysis, performed using the ldsc.py script, used LD scores derived from the European subset of the 1000 genomes-phase 3 reference panel. In accordance with best practices, GWAS summary statistics were cleaned ('munged') and harmonised to the common variants on the HapMap3 panel outside of the major histocompatibility region (MHC) The 'munging' step was performed using the R package GenomicSEM. These LDSR estimates were compared to those compared using LDSR with a slightly different variant weighting approach as implemented by R script Run_LCV.R (https://github.com/lukejoconnor/LCV).

#### Latent Causal Variable Models

Partial genetic causality is expressed as a posterior mean genetic causality proportion (GCP), whereby GCP > 0 implies partial genetic causality from the first trait to the second and *vice versa* given GCP < 0. We used the conventional threshold suggested by the authors of the LCV method (significantly non-zero |GCP| > 0.6) to infer partial genetic causality in either direction. It is important to note that the GCP is not an estimate of the magnitude of any potential causal effect but rather it establishes the direction in which a genetically inferred causal relationship may exist. In the case of a GCP value close to zero, this suggests that any observed genetic correlation probably arises due to horizontal pleiotropy, whereby there may be shared genetic risk, but there is no evidence that genetic liability to one of the traits influences the other. It is important to note that the LCV approach is not an estimate of the magnitude of any potential causal effect, but rather it establishes the direction in which a genetically inferred causal relationship may exist.

#### Mendelian randomisation – additional information

We investigated the effect of acne genetic liability on liability to psychiatric disorders, as well as the reverse model of psychiatric genetic liability to acne. MR is underpinned by the concept of using genetic 'proxies' [instrumental variables (IV)] of an exposure trait that are randomly inherited in the population under Mendel's laws, and as a result, the effect of the IV on the outcome trait of interest can be leveraged to estimate an exposure to outcome relationship. In addition to the CAUSE modelling, we also used some more traditional approaches that utilise a subset of variants strongly associated with the exposure ($P < 5\times10^{-8}$) as IVs, that is, the inverse variance weighted (IVW), penalized weighted median, weighted median, and weighted mode estimators. To select IVs, we used plink to clump genome-wide significant SNPs (--clump kb 1000 kb, --clump r2 <0.001).

#### Choice of beta prior for CAUSE

The default CAUSE implementation places a beta prior on *q* with asymmetric hyperparameters of one and ten - that is q ~ Beta(1,10). For exposure-outcome trait pairings with evidence of a more parsimonious fit of the 'causal' model, we also evaluated two other beta priors q ~ Beta(1,2) and q ~ Beta(1,100). The latter prior (1,100) places most of the prior weight on *q* values close to zero, and therefore, behaves in a more similar fashion to conventional MR approaches at the expense of higher false positive rates if the true extent of correlated pleiotropy is large. Conversely, the q ~ Beta(1,2) prior likely reduces power but is more robust to larger proportions of correlated pleiotropy. We evaluate all three modelling choices to ensure the robustness of favouring the 'causal model'. For trait-pairs where the sharing or causal model outperforms the null in terms of model fit, we also report the posterior median value of $q$ in the sharing model, which represents the proportion of variants that act through a ‘shared factor’.

**MR Clust Framework**

This approach allows instruments to be a member of a ‘null’ cluster, whereby the mean exposure-outcome effect is zero, and a ‘junk’ cluster unrelated to the others. Minimisation of the Bayesian Information Criterion was utilised to inform the number of clusters and cluster assignment. As a sensitivity analysis, we also examined the stability of inferred clusters by only assigning variants to a cluster with a conditional inclusion probability ≥ 80%.

**Interrogation of the PW-GWAS modelling**

The PW-GWAS model also outputs variant-level PPA where each variant is the causal one under each of the first three models. Using this, we computed 90% credible sets for the third model (shared causal variant in each region) in blocks outside of the MHC region with a PPA under the third model of at least 50%. Variants in these 90% credible sets were annotated to their closest gene using topr v.2.0.1. Credible sets were prioritised with a small number of consistent variants, as well as the genes with the highest maximum variant level PPA for the third model. We compared these findings to those of another Bayesian colocalisation approach (coloc v.5.2.3), which tests analogous hypotheses. PW-GWAS is an extension of coloc in the sense that it empirically estimates priors under each competing hypothesis, as opposed to providing these a priori with coloc, for which we used the default priors.

### SUPPLEMENTARY FIGURES

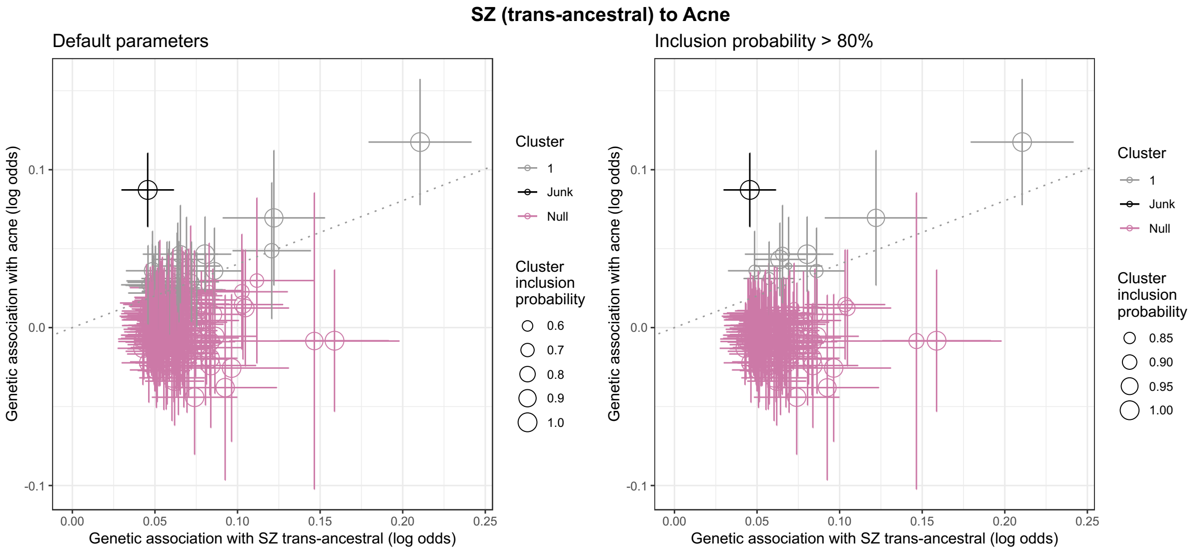


**Supplementary Figure 1. MR-clust results using the trans-ancestral schizophrenia GWAS**. The results of the MR-clust approach are visualised that investigated the association of individual schizophrenia IVs with acne. This is a mixture model framework that seeks to identify clusters of IVs with similar causal estimates. The size of the point denotes the cluster inclusion probability, relating to the conditional probability of cluster membership. The null cluster, coloured pink, relates to IVs with null effect, whilst the black “junk cluster” are variants that were not parsimoniously assigned to any cluster. The cluster of interest is denoted in grey, with a trend line indicative of the mean cluster effect. The leftmost panel utilised default parameters for cluster assignment, whilst the right panel only retained variants with an inclusion probability > 80%.

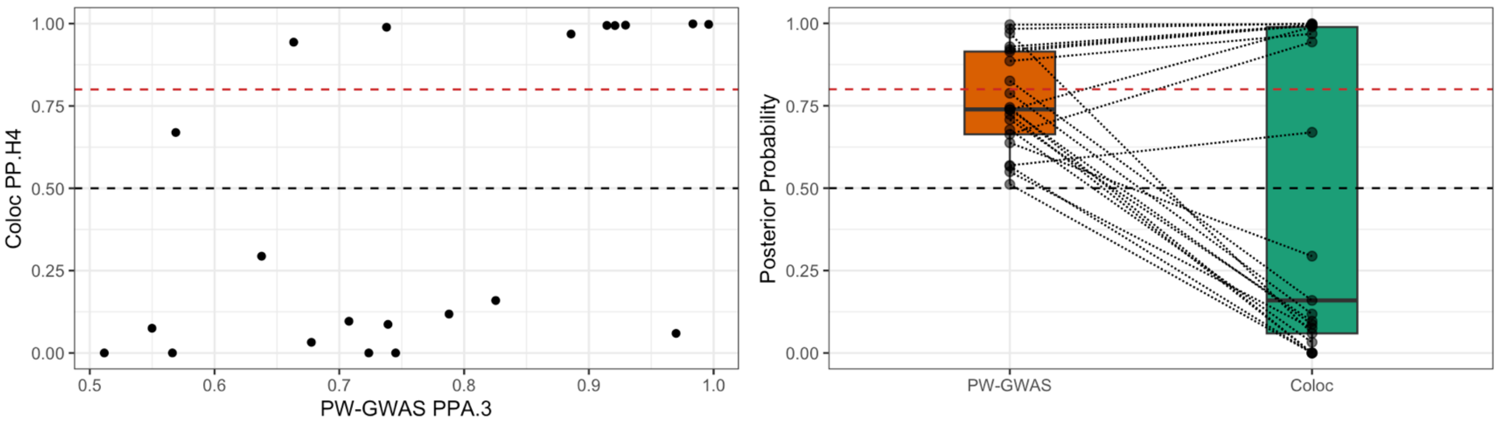


**Supplementary Figure 2. Comparison of results between two Bayesian colocalisation approaches (PW-GWAS and coloc)**. The right-hand panel x-axis denotes the posterior probability of the third PW-GWAS hypothesis (PPA.3), which is that the LD-block association with both traits driven by a single, shared causal variant. LD blocks with PPA.3 > 0.5 visualised. The y-axis denotes the equivalent coloc hypothesis (PP.H4). The left-hand panel directly visualises for each point its corresponding PW-GWAS PPA.3 versus coloc PP.4, with a box-and-whisker plot utilised to summarise the posterior probabilities for each method.

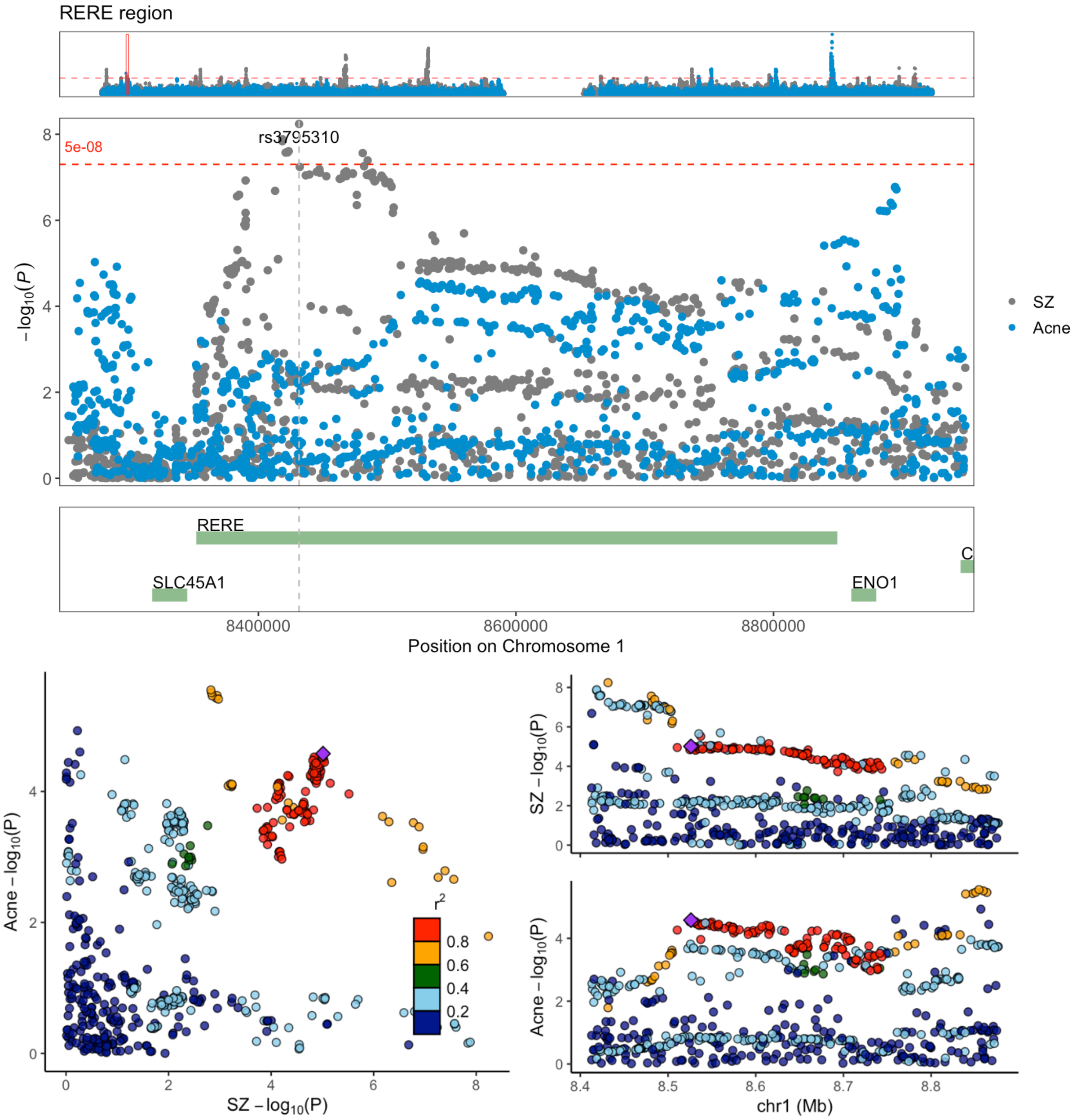


**Supplementary Figure 3. The association of the high-confidence schizophrenia risk gene *RERE* with acne.** The top panel visualises the *RERE* region in terms of the association of each variant with schizophrenia (grey) and acne (blue). The bottom panel directly compares the association signals in this locus between schizophrenia and acne.
